# Quantifying malaria acquired during travel and its role in malaria elimination on Bioko Island

**DOI:** 10.1101/2021.04.19.21255744

**Authors:** Daniel T Citron, Carlos A Guerra, Guillermo A García, Sean L Wu, Katherine E Battle, Harry S Gibson, David L Smith

## Abstract

**Background:** Malaria elimination is the goal for Bioko Island, Equatorial Guinea. Intensive interventions implemented since 2004 have reduced prevalence, but progress has stalled in recent years. A challenge for elimination has been malaria infections in residents acquired during travel to mainland Equatorial Guinea. We quantify how off-island contributes to remaining malaria prevalence on Bioko Island, and investigate the potential role of a pre-erythrocytic vaccine in making further progress towards elimination.

**Methods:** We simulated malaria transmission on Bioko Island using a model calibrated based on data from the Malaria Indicator Surveys (MIS) from 2015-2018, including detailed travel histories and malaria positivity by rapid-diagnostic tests (RDTs), as well as geospatial estimates of malaria prevalence. Mosquito population density was adjusted to fit local transmission, conditional on importation rates under current levels of control and within-island mobility. We evaluated the impact of two pre-erythrocytic vaccine distribution strategies: mass treat and vaccinate, and prophylactic vaccination for off-island travelers. We propagated uncertainty through the model through an ensemble of simulations fit to the Bayesian joint posterior probability distribution of the geospatial prevalence estimates.

**Results:** The simulations suggest that in Malabo, an urban city containing 80% of the population, there are some pockets of residual transmission, but a large proportion of prevalence is attributable to malaria importation by travelers. Outside of Malabo, prevalence was mainly attributable to local transmission. We assess the uncertainty in the local transmission vs. importation to be lowest within Malabo and highest outside. Using a pre-erythrocytic vaccine to protect travelers would have larger benefits than using the vaccine to protect residents of Bioko Island from local transmission. In simulations, mass treatment and vaccination had short-lived benefits, as malaria prevalence returned to current levels as the vaccine’s efficacy waned. Prophylactic vaccination of travelers resulted in longer-lasting reductions in prevalence. These projections were robust to underlying uncertainty in prevalence estimates.

**Conclusions:** The modeled outcomes suggest that the volume of malaria cases imported from the mainland is a partial driver of continued endemic malaria on Bioko Island, and that continued elimination efforts on must account for human travel activity.

## Background

Importation of malaria represents an important barrier to elimination in many cases. Indeed, there are many known settings in which human travelers contribute to outbreaks when they bring malaria parasites from a high-transmission setting to a low-endemic or pre-elimination setting [1, 2, 3, 4]. It is important that malaria elimination programs which operate in such settings understand the risk of reintroduction of malaria parasites by visitors or by residents who travel away from home and return with infections [5, 6, 7, 8].

Bioko Island in Equatorial Guinea represents a setting where malaria endemicity has been reduced relative to neighboring areas in the region, and where people travel frequently. The Bioko Island Malaria Control Project (BIMCP) began in 2004, implementing an extensive program of indoor insecticide spraying, longlasting insecticidal net distribution, expanding access to diagnostics and treatment, and surveillance. The program was later renamed as the Bioko Island Malaria Elimination Project (BIMEP), reflecting the ambition to interrupt malaria transmission on Bioko Island altogether. As of 2015, the average malaria parasite rate (*Pf* PR) in children 2-14 years old has fallen from 0.43 to 0.11 [9]. At the same time, there has been a sharp reduction in the viable vector population on the island [10, 11, 12]. Despite reductions in transmission, malaria persists across Bioko Island. Many areas remain receptive to malaria outbreaks, as was documented in Riaba District in 2019 [13]. The changes brought about through the BIMEP represent tremendous progress, yet that progress has stagnated in recent years and prevalence has not decreased further despite ongoing efforts to contribute to elimination [9, 14].

One hypothesis for why malaria persists in certain areas of Bioko Island is that cases may be attributable to travelers to mainland Equatorial Guinea [9]. While *Pf* PR has decreased on Bioko Island since 2004, there has not been the same concerted effort to reduce malaria burden in mainland Equatorial Guinea and prevalence remains high in that region [15], estimated as 0.46 among all age groups in a recent study [16]. One study of children found that those who reported recent travel to the mainland were much more likely to be infected than those who had not (56% vs 26% in 2013; 42% vs 18% in 2014) [6]. The same study also found that areas with high proportion of travelers increased the risk of malaria infection in non-travelers [6]. A subsequent analysis of island-wide surveillance data produced geospatial estimates of malaria prevalence across Bioko Island, and found significantly higher prevalence among travelers to mainland Equatorial Guinea [17]. The data collection efforts through BIMEP have resulted in a detailed understanding of the current state of malaria transmission on Bioko Island. The next step, however, is to assess options for where and how to intervene against the disease. In this setting, the BIMEP needs to understand why it is that progress has slowed and what changes that could be made to the malaria intervention program are most likely to continue reducing the case burden on Bioko Island. To this end, we extend the analysis of [17] using a simulation model to represent transmission patterns across Bioko Island, to further quantify the fraction of cases attributable to off-island travel, and lastly to predict the impact of possible changes to the current set of interventions.

We approach this problem using a simulation model of malaria transmission patterns on Bioko Island. We construct the model to reflect our current best understanding of the epidemiological ground truth from 2015-2018. We then use the model to quantify the fraction of cases which may be attributable either to local transmission or to exposure while traveling. That is to say, we are able to map where it is that people experience exposure risk, providing important and actionable information for how and where to intervene. We also are able to use the model to quantify the efficacy and impact of potential future interventions. Specifically, we investigate the potential impact of distributing a vaccine which confers preerythrocytic protection against malaria to see whether such an intervention could result in eliminating malaria in the long term [18].

The structure of the paper is as follows: we will begin by describing the data sources which we use to understand the transmission environment on Bioko Island. We then describe our simulation model and how we used our data sources to calibrate the model — this calibration step is important, because it allows us to derive local exposure risk from local estimates of prevalence. Knowing the local exposure risk, we are then able to use the simulation to differentiate between cases resulting from local transmission and cases acquired during travel. Overall, we are able to quantify the importance of travel and importations in the transmission setting of Bioko Island, and show that there are portions of the island where prevalence is sustained through imported cases. Lastly, we simulate the impact of expanding the current intervention package and deploying a vaccine against malaria.

## Methods

### Data sources

Much of the data which we use to design and calibrate our simulation model of malaria transmission on Bioko Island was collected by the BIMEP and the National Malaria Control Program (NMCP). Population census data were collected as part of two campaigns for distributing long-lasting insecticidal nets across the island in 2015 and 2018 (Figure 1b) [19, 20, 21]. Although the bednet campaigns only visited 88% of households on the island and hence underestimated the overall population, we rely on these data as they represent the most accurate map of human population on the island. Since the beginning of malaria control on Bioko Island in 2004, the BIMEP has performed extensive annual malaria indicator surveys (MIS) that have collected epidemiological, demographic, and socioeconomic data. We draw upon MIS data collected in each of the years from 2015 to 2018, which sampled an average of 16,500 respondents each year, to analyze transmission patterns on Bioko Island [22, 23, 24, 25]. MIS data were again collected in 2019 and 2020, but the analyses in the present manuscript were performed before those data were available.

**Figure 1.**
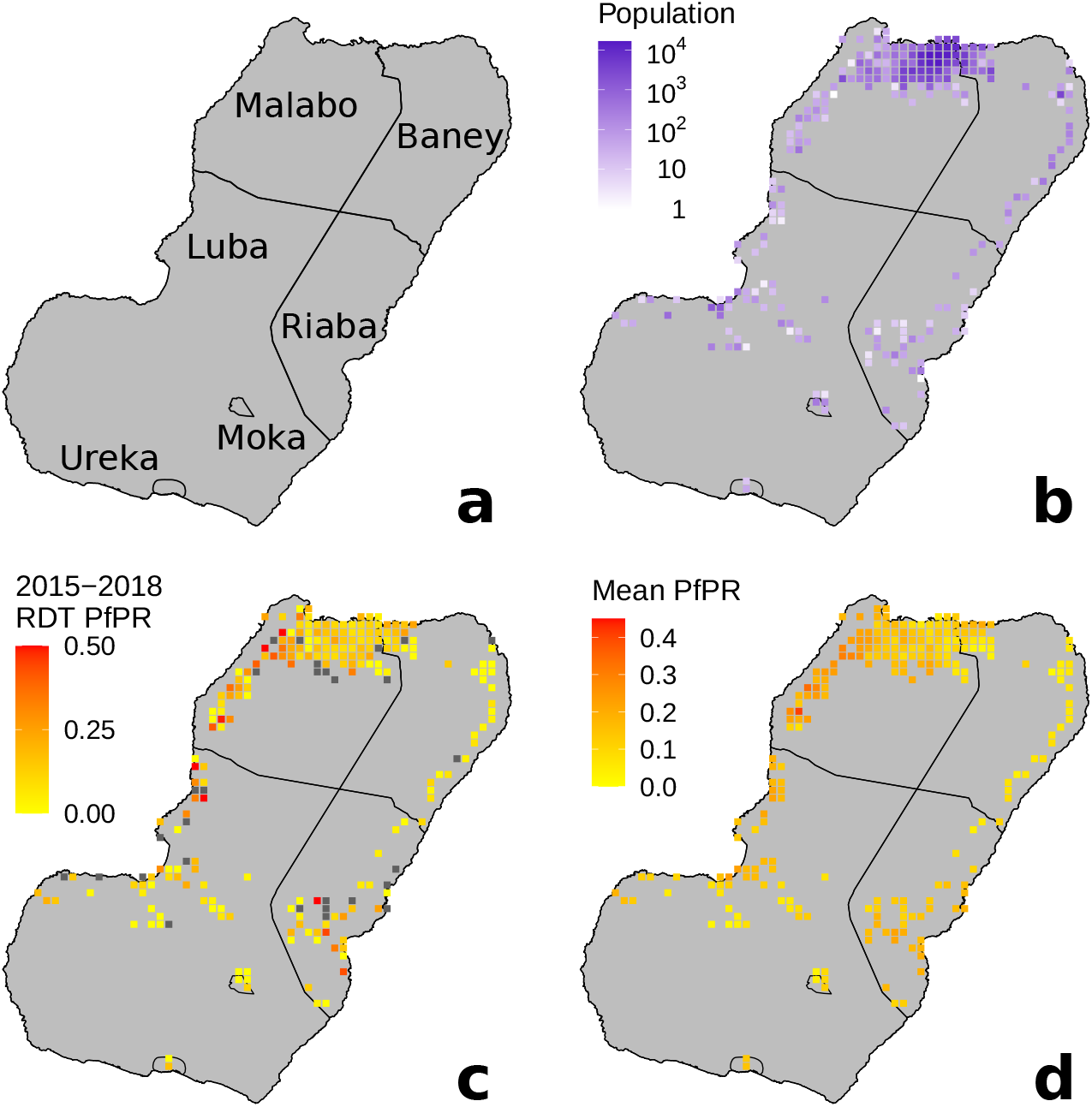
Maps of Bioko Island. All squares represent 1km × 1km areas. **a** Map of administrative units on Bioko Island. **b** 2018 population map. **c** Parasite rate *Pf* PR ascertained through the use of rapid diagnostic tests as part of the 2018 MIS. **d** Mean geostatistical estimates of *Pf* PR, reproduced with permission from [17].

The MIS data include results from rapid diagnostic tests (RDT; CareStart Malaria Pf/PAN (HRP2/pLDH) Ag Combo RDT, AccessBio Inc, Monmouth, USA), used to diagnose survey participants for the presence of detectable *Plasmodium falciparum* parasites (Figure 1c). The RDT results in the MIS data make it possible to map the spatial distribution of occurrences of malaria infection. Guerra et al. utilized the RDT data to produce geostatistical estimates of *Plasmodium falciparum* parasite rate (*Pf* PR) [17]. The geostatistical estimation techniques use the RDT results together with geospatial environmental covariates to estimate malaria prevalence across the island, accounting for the uneven distribution of samples taken by the survey [26]. These estimates allow us to see how the prevalence varies across geographical space, with the highest prevalence on the island occurring along the northwest coast and in the southeast; a reduced prevalence in the high population density, urbanized areas in the capital city of Malabo; and reduced prevalence in the elevated regions nearer to the center of the island. The overall mean estimated *Pf* PR is around 0.12 for the entire island. Note that the *Pf* PR estimates reflect the current level of interventions on the island. The *Pf* PR estimates are made for all age groups, as there were no significant differences between *Pf* PR measurements made for children (ages 2-10 years) and the overall sample population [17]. The median surface of *Pf* PR estimates are shown in Figure 1d, reproduced with permission from [17].

The MIS asks respondents about recent travel history, providing information on where it was that people could have become exposed. For the MIS in 2015- 2017, respondents reported whether they had recently taken a trip where they had spent at least one night away from their home residence in the preceding 8 weeks. Respondents who reported at least one trip also reported one of seven possible destinations, to each of the regions of Malabo, Baney, Luba, Riaba, Moka, Ureka, or off-island (Figure 1a). Most of the travelers (84%) reporting off-island travel reported going to mainland Equatorial Guinea [17]. The 2018 MIS included an additional question on travel duration, such that we could assess how many days travelers had been away from home [27].

Given that so many travelers reported going to mainland Equatorial Guinea, our model also required knowing estimates of prevalence in that region. We draw the overall prevalence in the region of mainland Equatorial Guinea from the Malaria Atlas Project, which estimated a median *Pf* PR of 0.43 in children aged 2-10 at the time of MIS data collection [15]. This estimate is corroborated by a study conducted in 2015 by Ncogo et al., which reported an overall *Pf* PR in the region to be 0.46 [16] among all age groups. The Ncogo et al. study did find geographical variation in *Pf* PR, with elevated prevalence as high as 0.58 in rural areas and 0.34 in urban areas [16], but the MIS travel data do not contain enough detail to report exactly where travelers spent their time in the mainland. For this reason, along with the fact that even in lower-prevalence areas the *Pf* PR remains far higher than on Bioko Island, we use the median *Pf* PR estimate for the mainland region. That the prevalence is so much higher on the mainland than on Bioko Island suggests that travelers would experience a much higher risk of infection while visiting the mainland than they would at home. This is consistent with the prior studies and analyses performed which suggest that there is an elevated risk of prevalence among those who had recently traveled to mainland Equatorial Guinea [6, 9, 17].

Lastly, the MIS data also included some basic information on treatment-seeking behavior. Respondents reported whether they had recently experienced a fever and whether they sought treatment for malaria.

### Designing the simulation model

We constructed a family of models to simulate malaria transmission patterns on Bioko Island, based on the census and epidemiological data collected from 2015-2018. The models are continuous-time, event-driven agent-based stochastic simulations that describe how individual human hosts become infected through contact with mosquito vectors in the environment; how malaria infections run their course; and how infected individuals contribute to onward transmission of subsequent infections. The models are spatially explicit, meaning that prevalence and local transmission intensity are allowed to vary across geographical space. The model also simulates human travel behavior, and human hosts may experience different levels of transmission risk as they move from one location to another. Parameters describing local transmission and human mobility were fit to be consistent with the geospatial analysis of prevalence and MIS data (see below). Code supporting the simulations may be found in the `macro.pfsi` directory at https://github.com/dtcitron/bioko_island_travel_materials, and an explanation for how the simulation program works may be found in Section 1 of the Supplementary Information.

Within the simulation, the resident population of Bioko Island is based on the 2018 census data, which were aggregated into 241 gridded 1km×1km map-areas [21] (as shown in Figure 1b). Each populated map-area from the census is represented as an isolated patch in the simulation, home to as many human hosts as in the corresponding map-area from the census data. Patch residents spend most of their time in their home patches but occasionally travel to other ones. To simulate malaria importation, we also include a 242nd patch to represent the destination of off-island travel (mainland Equatorial Guinea). This final patch serves as a boundary condition for the simulation: Bioko Island residents import cases through experiencing exposure risk while traveling off-island, and the 242nd patch represents the off-island transmission environment.

The core of the simulation model represents transmission of malaria parasites between human hosts and the vectors. We base the core of our transmission model on the Ross-Macdonald model, which includes a modeled description of human hosts, the population dynamics of the vector population, how the vectors and humans interact with one another, and the course of infection in a malaria-afflicted individual [28, 29].

Figure 2 shows a simplified schematic representing the modeled compartments for both mosquitoes and humans within a single patch. The model tracks mosquito population dynamics: within each patch, adult mosquitoes emerge at a rate which reflects the local ecology and die at a constant rate. Adult mosquitoes become infected with malaria parasites when they blood feed on infectious human hosts. After the extrinsic incubation period, surviving adult mosquitoes then become infectious. Each day, within each patch we generate a number of infectious bites based on local population of infectious mosquitoes and distribute those infectious bites across the human hosts who are present that day. Thus, susceptible human hosts can become infected. Infected human hosts may develop symptoms and consequently seek treatment, at which point their infections become cleared and they remain protected against new infections for a short period. Two final model features are not shown in Figure 2. The complete transmission model simulates together many patches, and allows for human hosts to travel between them. The simulation model also has the capability to simulate the distribution of a pre-erythrocytic vaccine, where vaccinated people experience a lower infection risk.

**Figure 2.**
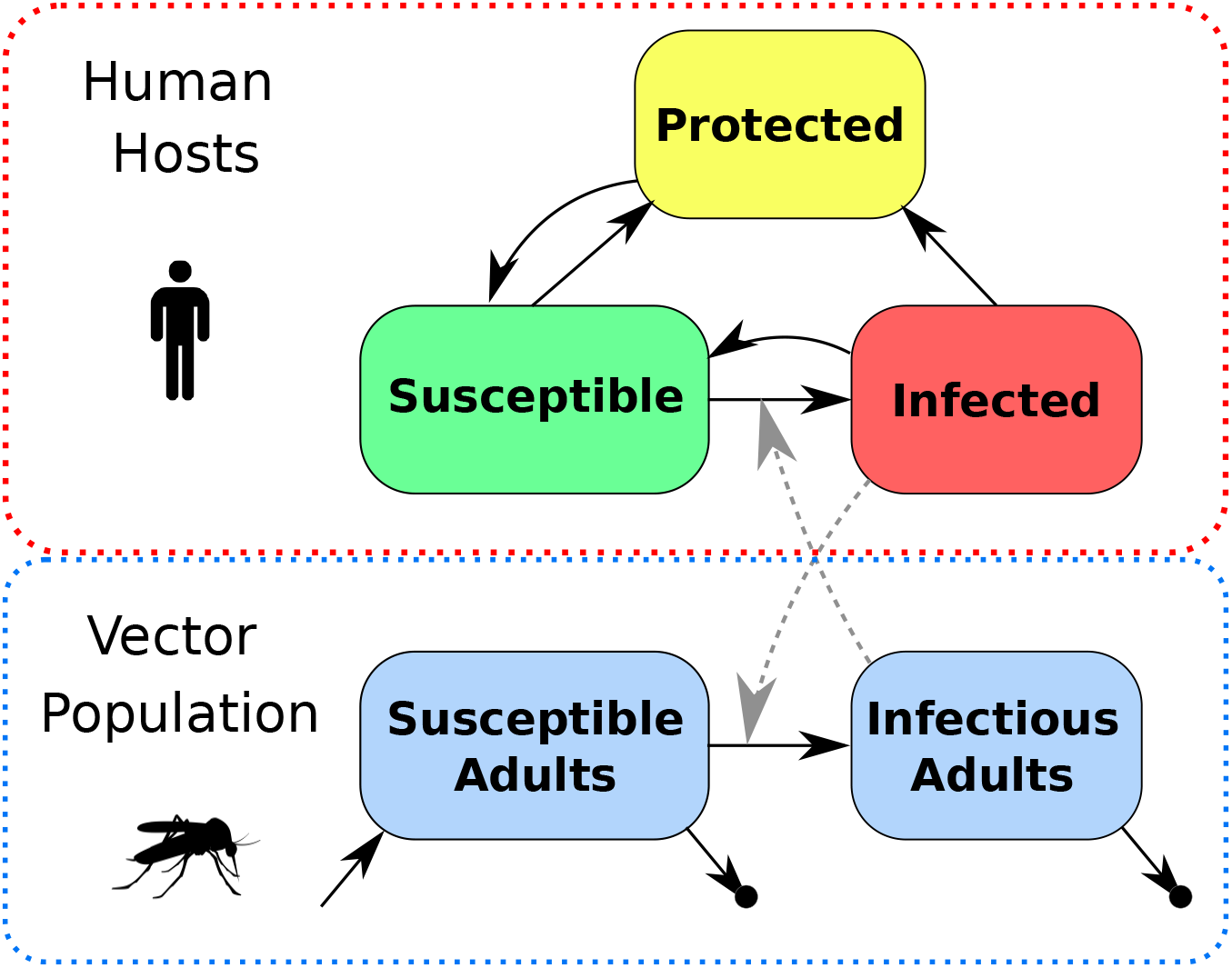
Diagram of Simulation Model. Human hosts begin in a Susceptible state and become Infected through contact with infectious mosquitoes. Infected human hosts may develop symptoms and seek treatment, which clears the infection and moves them to a Protected state where they resist infection for 30 days. Susceptible human hosts may also enter the Protected state if they are given anti-malarial prophylaxis. Adult mosquitoes emerge at a constant rate and become infected through contact with infectious human hosts, and then become infectious after surviving through the extrinsic incubation period. Mosquitoes die at a constant rate. Not shown are human hosts moving between different patches and vaccination.

### Calibrating the simulation model

The key to calibrating each simulation model is to set the mosquito population density such that it sustains a level of transmission intensity which subsequently produces the correct *Pf* PR within each patch. We calibrate the simulation using a geospatial estimate of *Pf* PR within each patch drawn from the Bayesian posterior probability distribution [15, 17], and then use our transmission model to translate *Pf* PR into a local force of infection (FOI). Each patch’s FOI is a quantitative representation of the transmission risk experienced by human hosts found there. Knowing the FOI, we again use our transmission model to derive that patch’s mosquito population density. Within the simulation, the mosquito population density produces blood feeding activity, which in turn affects the transmission risk (FOI) which finally gives rise to the geospatial *Pf* PR estimates when the mosquito and human populations are coupled together within the simulation. Thus, we are able to calibrate the vector ecology across the different patches on the island such that we reproduce the geospatial *Pf* PR estimates.

Knowing *Pf* PR is only the first step to calibrating the simulation model. We initialize the simulation by setting the the mosquito population density in each patch. The simulation translates each patch’s mosquito population density into local FOI. As a consequence of the malaria transmission and infection model, the simulation translates FOI into *Pf* PR. The trick to constructing the simulation model is to calibrate the mosquito population density and FOI in each patch such that the resulting *Pf* PR matches the mapped *Pf* PR estimates from data. We adapt this procedure from the source-sink analysis described in [30, 31] and describe it in detail in Section 2 of the Supplementary Information. From the perspective of an individual human host, their risk of becoming infected with malaria is the result of the FOI they experience. The average FOI is computed as a sum of the FOI experienced in each patch visited by that individual weighted by the duration of time spent in each of those patches.

### Incorporating Travel Data

Accounting for human hosts’ travel patterns is the last step required for properly calibrating FOI. The total FOI experienced by an individual host includes both FOI at home as well as FOI experienced while traveling away from home. For this, we construct a model of human travel patterns on Bioko Island and parameterize that model using the MIS travel data [27]. We model each human host’s travel behavior in three steps: the human host chooses when to leave their home patch; chooses the destination; and chooses how many days they spend away before returning to their home patch. Within the simulation, each individual requires a set of parameters that includes the frequency of travel; the multinomial probability distribution for determining travel destination; and the mean duration of their trip [32]. This model of movement behavior is too simple to allow for trips with multiple destinations, but it is nevertheless consistent with the MIS travel history data. We estimate the frequency of leaving home based on the frequency of trips reported by MIS respondents. We estimate the probability of traveling to each travel destination based on the relative frequencies of trips from each patch to each destination region. We estimate the duration of trips based on the distribution of trip duration reported — an average of about 10.5 days for travel within Bioko Island and 20 days for travel to mainland Equatorial Guinea. Data for fitting travel frequencies and probabilities were available from the MIS from 2015-2018, but data for fitting travel duration were only available with the most recent 2018 MIS [27]. Refer to Section 3 in the Supplementary Information for a detailed description of how we parameterized the movement model. For simplicity, all human hosts from each patch have the same set of movement parameters, but we allow those parameters to vary from patch to patch according to the MIS data. For example, individuals who live in the southern parts of Bioko Island travel more frequently and tend to choose Malabo as the destination, whereas individuals who live in Malabo travel less frequently but tend to choose mainland Equatorial Guinea as the destination. We assume that we can ignore migration of mosquitoes between patches.

### Estimating uncertainty

One of the benefits of using a simulation model is that it enables us to produce results which reflect the uncertainties underlying the data used to calibrate the model. The geostatistical *Pf* PR estimates include mean surfaces across the island (plotted in Figure 1d) as well as a full ensemble of draws from the joint posterior distribution. Each draw from the joint posterior distribution by itself represents a map of *Pf* PR estimates across Bioko Island that also accounts for spatial correlations between the *Pf* PR in different patches. The full ensemble of draws from the joint posterior distribution represents a sample which reflects the uncertainty of the mapped *Pf* PR estimates [15]. Using the ensemble of draws, we construct an ensemble of simulated outcomes. For each scenario that we simulate, we create 1000 simulation runs. Each simulation run is calibrated using its own *Pf* PR surface draw. The full ensemble of simulation runs represents a family of models whose outputs reflect the variability in simulated outcomes when considered together. We can express these variations in terms of error bars or confidence intervals. The error bars shown in the subsequent plots reflect both the uncertainty inherent to the stochastic simulation as well as the uncertainty associated with the *Pf* PR maps.

## Results

### Infections attributable to off-island travel

The fully calibrated simulation model now enables investigations of how the volume of human travel from Bioko Island to the mainland influences the transmission patterns found in the MIS data. We distinguish the infections that are caused by local transmission on Bioko Island from the infections which affect travelers on the mainland. We define the travel fraction as the fraction of prevalence which is attributable to off-island travel. This can be measured within the simulation by eliminating exposure risk at home, setting the local FOI to zero on Bioko Island in the simulation. Figure 3a is a map of this travel fraction, showing that for many areas in Malabo and a few areas in the Luba district in the south a high fraction of the estimated prevalence may be attributable to travel. This result is consistent with the entomological surveillance data, which have shown a significant decrease in viable vector populations since the BIMCP began [10, 11, 12]. While more thorough entomological monitoring is still required to characterize the transmission ecology in Malabo and elsewhere on the island in greater detail, it is plausible that the apparently diminished vector populations in the city are not solely responsible for sustaining the high malaria prevalence in the area.

**Figure 3.**
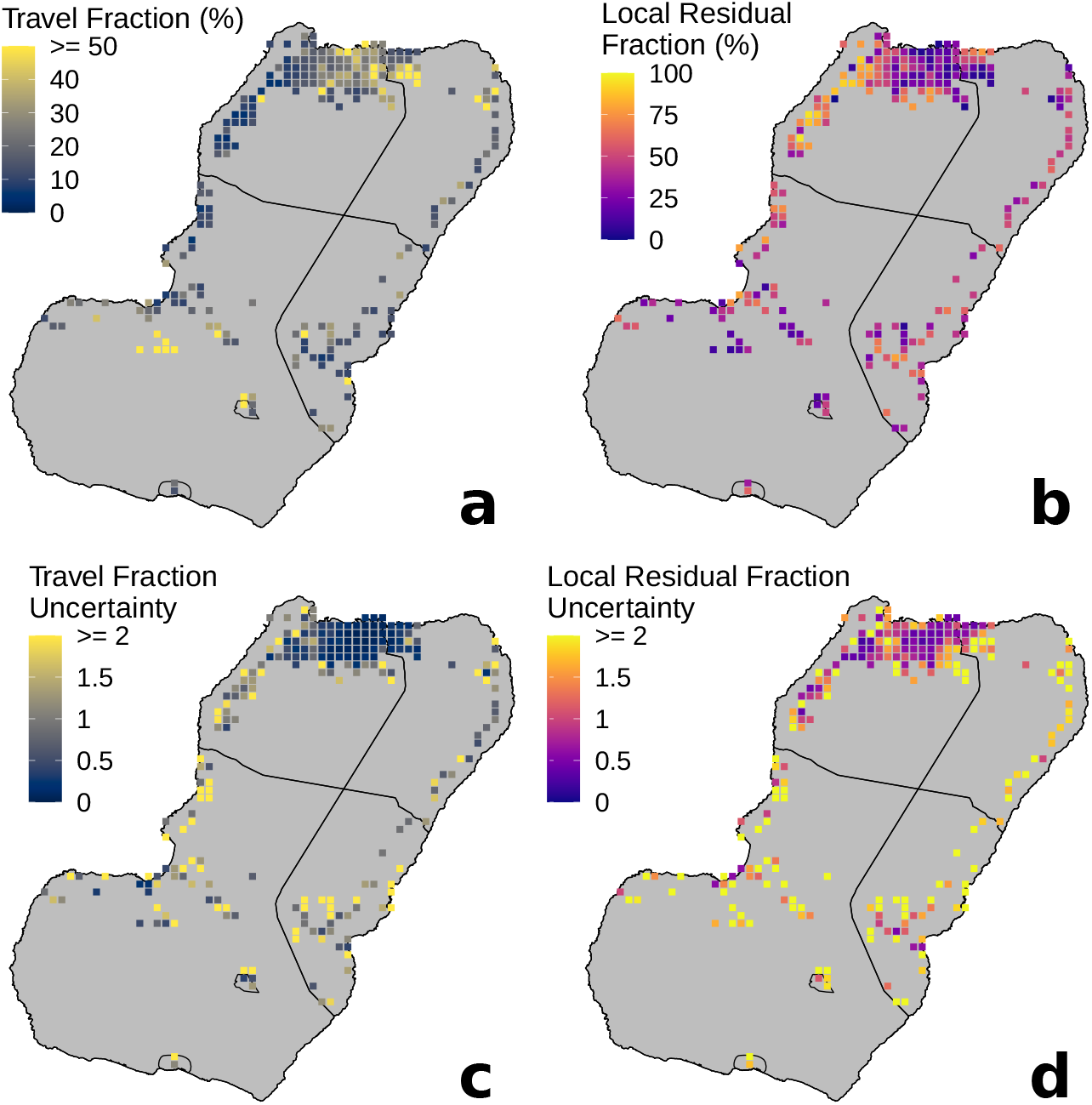
Travel Fraction and Local Residual Transmission. **a** Travel fraction, or the fraction of *Pf* PR attributable to infections contracted while traveling to the mainland. **b** Local residual fraction, or the fraction of *Pf* PR attributable to local transmission. **c** Travel fraction uncertainty: the standard deviation of the simulation results, divided by the mean of the simulation results. (*σ*_*T F*_ */µ*_*T F*_) **d** Local residual fraction uncertainty. (*σ*_*LRF*_ */µ*_*LRF*_)

Similarly, we define the local residual fraction as the fraction of prevalence which is attributable to local transmission only. This can be measured within the simulation by eliminating the exposure risk to travelers and setting FOI to zero off-island while preserving local transmission patterns. Figure 3b is a map of this travel fraction, showing that once the influence of off-island transmission is removed, there are some areas particularly along the northwest coast where local transmission is still responsible for many malaria cases.

These maps make it possible to discern where different types of interventions might be most effective. Deploying additional interventions against the vector — such as indoor residual spraying, long-lasting insecticidal nets, and larval source management — in areas where the Local Residual Fraction is low is unlikely to lead to a strong measurable decrease in *Pf* PR. Instead, the areas with high local residual fraction likely have a malaria burden which is driven by local transmission. By the same token, a different set of interventions is likely necessary to further reduce *Pf* PR in Malabo where the majority of annual malaria cases occur.

The modeling methodology also makes it possible to evaluate the uncertainty associated with the modeled outcomes across different parts of the island. The ensemble of simulated results show variation that reflects the underlying uncertainty in the *Pf* PR estimates used to calibrate the simulation as well as the uncertainty from the stochastic simulation. Figures 3a and b map the ensemble means of travel fraction and local residual fraction, respectively. Figures 3c and d map the uncertainty in the travel fraction and local residual fraction, respectively, where uncertainty is the ensemble standard deviation divided by the ensemble mean (*σ/µ*, or coefficient of variation), showing the characteristic variations that appear across the ensemble of simulation runs. For both maps, we see lower uncertainty in the densely populated areas in and around Malabo in the north and higher uncertainty in the less populated areas elsewhere on Bioko Island. The interpretation here is that we can be most confident about the estimates of travel fraction and local residual fraction in and around Malabo than in other areas of the island.

### Simulating additional interventions

We next use the simulation to estimate the impact of expanding the program of interventions. It has been suggested that a vaccine may be effective for further reducing the number of malaria cases on Bioko Island [33]. Already there have been vaccine efficacy studies on Bioko Island of a pre-erythrocytic vaccine which works to block the maturation of parasites prior to blood stage infection [18]. Within the simulation, this works by reducing the probability that an infectious bite causes a new infection in a human host by 50% for an average of 10 months (sampling from a normal distribution with mean duration of effect 300 days *±* 30 days standard deviation), providing temporary protection for a little less than a year. The parameters used here are largely hypothetical, although we find that the results reported below are robust to changes in the parameters (unless the vaccine is assumed to be indefinitely and 100% effective in blocking new infections). The vaccine in the simulation is administered along with parasite-clearing treatment, so human hosts who receive a vaccine lose their infections if they were infected and enter a protected state for a short period. We assume that travelers account for any latent periods such that the vaccine’s protection is in effect by the time they leave the island. We simulate three possible scenarios: administering vaccines to all travelers who leave the island; vaccinating everyone on the island; and reducing the risk of infection among travelers by reducing the transmission risk in mainland Equatorial Guinea by 50%.

Figure 4 shows the simulated future trends of malaria prevalence in each of these scenarios for an area in urban Malabo (a, upper row) and a more rural area in Riaba in the south (b, lower row). For each scenario we simulate 1000 simulation runs. The black lines represent the mean of the ensemble of simulation runs and the error bars represent one standard deviation above and below the mean. We choose to show the error bars to explicitly illustrate the uncertainty in our simulation results: The error bars reflect both the uncertainty inherent to the stochastic simulation as well as the uncertainty underlying the *Pf* PR estimates.

**Figure 4.**
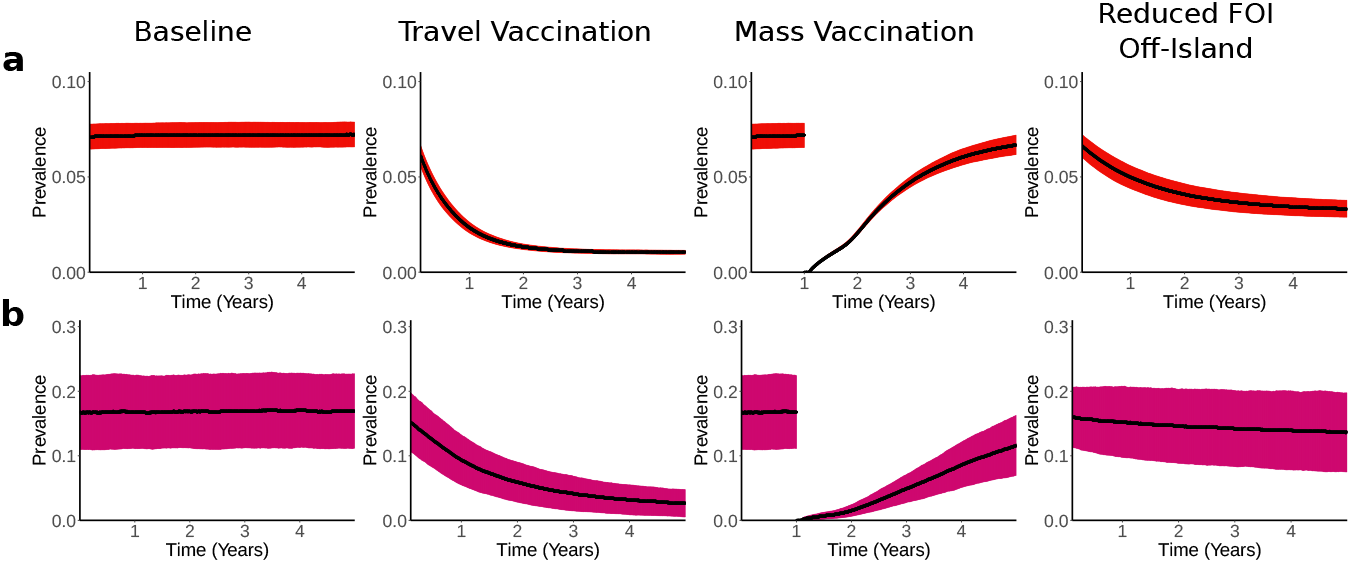
Simulated Impact of Additional Interventions. Showing baseline case and three additional intervention scenarios for an area in urban Malabo (**a**, top row) and Riaba in the South (**b**, lower row). The black lines represent the mean result across 1000 simulation runs and the red error bars represent the mean *±* one standard deviation.

The leftmost column in Figure 4 shows the baseline case, of how *Pf* PR would persist if we assume that the island’s transmission ecology does not change and the BIMEP makes not changes to its current suite of interventions. (This scenario does not reflect alternative vector control measures currently being considered.) The second column from the left shows the impact of travel vaccination: an individual receives a vaccine dose the first time each year that they schedule a trip off-island. Administering treatment and vaccination to travelers to the mainland once per year results in a strong, steady decrease in prevalence over time. The third column shows the impact of distributing the vaccine to all residents on the island. From the mass treatment, this would very quickly reduce the prevalence to zero, but as the protective effects of the vaccine begin to wear off we begin to see that prevalence increases back to the baseline level after a few years. Indeed, the influence of off-island transmission is strong enough that even if the parasite population is reduced to zero on the island it is likely to be replenished after a few years through contact with the mainland. A single round of mass vaccination is unlikely to halt transmission for very long, meaning that multiple subsequent rounds of mass vaccination would be required to sustain any temporary progress and provide lasting protection for the island’s population. We emphasize that the simulated vaccine deployment represents a best-case scenario, and in practice it is more likely that the vaccine would reach the population over a period of many months or years through a sustained distribution campaign. Varying the parameters in the simulation that encode the vaccine’s efficacy and duration of protection does not qualitatively change the conclusions reported here: as long as the vaccine’s effect wears off after a short period, mass vaccination will not result in a reduction in malaria prevalence much longer than the vaccine’s duration on Bioko Island, as the volume of imported cases will remain too high.

Directly comparing the travel vaccination and the full-island vaccine deployment strategies, the travel vaccination requires fewer resources for distribution but also requires more doses of vaccine if the plan is to vaccinate all travelers over the course of many years. Within the simulation, over 240,000 doses of vaccine are needed to vaccinate all island residents. This number is likely an underestimate: the population counts used to create the simulations came from a 2018 bednet distribution campaign which did not reach all households, and the island’s total population has been estimated to be as high as 335,000 by the government of Equatorial Guinea. On the other hand, distributing vaccines to travelers only would require on average fewer than 130,000 doses each year. This number is likely an overestimate, seeing as many who travel off-island do so frequently while many others do not. The simulation does not explicitly account for this heterogeneity across different individuals, hence only requiring vaccinations for travelers may result in reducing the required volume of vaccine doses.

The rightmost column of Figure 4 shows the impact of reducing the FOI on the mainland by 50% through an intervention program similar to the one run by BIMEP on Bioko Island. There is a significant reduction in *Pf* PR in the areas with high travel fraction (a, upper row), but less of a reduction in areas with higher local residual fraction (b, lower row). Thus, this change would not result in a strong decrease in malaria burden everywhere on Bioko Island. Furthermore, this scenario is largely hypothetical, as it would require a major investment of resources and time to achieve these changes on the mainland, but it serves as a comparison with the vaccine-related interventions.

## Discussion

Our analysis demonstrates the important role of imported malaria in sustaining the reservoir of malaria parasites on Bioko Island, Equatorial Guinea. The malaria parasite populations of Bioko Island in general, and urban Malabo in particular, are well-connected to the parasite populations of mainland Equatorial Guinea. The analysis suggests that there are many areas, particularly in urban Malabo, where most malaria cases are likely attributable to infections contracted while traveling off-island. There are also areas outside of Malabo where residual local transmission appears to be high enough to sustain endemic transmission, with some malaria being due to onward transmission from imported cases by residual mosquito populations. The current distribution of malaria on the island is highly heterogeneous, but there is substantial uncertainty about where the last residual transmission foci remain, in part, because their locations are masked by high rates of imported malaria. Malaria importation thus represents one of the most important challenges to malaria elimination on Bioko Island.

While a pre-erythrocytic vaccine would improve the prospects of elimination on Bioko Island, we show that the most effective use of a pre-erythrocytic vaccine would be to prevent infections in travelers, which would also have a large effect on reducing the number of malaria cases imported to Bioko Island. For most travelers, intensive vector control measures in place across the island mean that the risk of malaria is much higher while traveling. A travel vaccine would thus have a direct benefit for those who travel. These simulations suggest that high coverage with a highly effective travel vaccine would also have sustained effects on malaria prevalence in Bioko Island, effectively increasing its isolation from the mainland. The simulations suggest that administering vaccines directly to travelers is more effective at lowering overall *Pf* PR in the long term than vaccinating everyone on the island all at once. As long as importation is halted or slowed, mass vaccination would likely require multiple rounds across many years in order to protect the island population over the long term. While acknowledging operational challenges, distributing the vaccine to travelers is likely to require fewer resources than distributing the vaccine to all island residents, given only two points of departure for leaving the island by sea or air. Broadly speaking, the vaccine discussed here is largely theoretical and any further analysis for discussing a specific vaccine would require parameterizing according to clinical trial data [18]. Operational concerns would need to account for the duration of a vaccine’s protective efficacy and delays when multiple vaccine doses are required for protection. Simulations show that mass treatment and vaccination of travelers on Bioko Island would not be as effective because of the short-lived efficacy of the vaccine, and because the reservoir of parasites would be rapidly renewed by imported malaria.

In using simulation models to evaluate potential policy scenarios, we recognize the importance of quantifying and propagating uncertainty. In this study, we have shown that our recommendations are robust to statistical uncertainty in the spatial distribution of malaria prevalence. The simulation model is spatially explicit, where mosquito populations and transmission intensity are allowed to vary across different locations. In each population, the mosquito populations were fit to a different modeled surface selected at random from the Bayesian posterior probability distribution (cite: joint simulation). The policy recommendation is robust to spatial uncertainty in that it considers reasonable alternative formulations of a model; the policy implications are consistent across the full range of uncertainty in the underlying MIS prevalence data. We acknowledge that there are other important sources of uncertainty to be considered. Our modeling framework, which draws from several different data sets, could be extended to perform a full sensitivity analysis across many different data inputs, and illustrate for stakeholders how different data sources contribute to overall uncertainty.

An important concern for malaria is uncertainty about the future, such as the environmental conditions that affect mosquito populations, the malaria interventions that suppress transmission, and connectivity to the mainland. In particular, our simulations assume that interventions currently used by BIMEP, such as bed net distribution and indoor residual spraying, would remain in place and continue to be at least as effective as they currently are. The assumptions about local transmission vs. imported malaria describe mosquito ecology and malaria transmission using MIS data from the years 2015-2018. Unfortunately, there is evidence that the transmission setting has changed over the last two years. In 2019, there was a marked increase in local transmission and cases in several areas including the northwestern coast and in Riaba District in the southeast [13], likely due to changes in the local ecology. Two important factors were higher than normal rainfall and construction projects that created mosquito habitats. We did not explicitly consider these scenarios. In 2020 the BIMEP expanded their use of indoor residual spraying in Malabo as a response to evidence of increased EIR in the area, but it seems likely mass treatment and vaccination would, perhaps, be one way of responding to such perturbations.

Additionally, much of the conclusions suggested through our modeling methodology follow from the presence of a high volume of travelers between Bioko Island and the mainland. As of 2020, much of the volume of travelers has been reduced due to port closures implemented as a response to the COVID-19 pandemic. Our modeling results would suggest that dramatically reducing the number of travelers between the mainland and Bioko Island for a sustained period of many months might result in a reduction in prevalence in areas where travel fraction is high. At the same time, we might expect to see no such decreases in locations on the island with high local residual transmission. This represents a natural experiment affecting malaria at the same time as the ecological changes mentioned above; as MIS data from 2020 become available, it may be possible to support evidence for the connections between travel and malaria cases on Bioko Island if cases do begin to decrease.

Some aspects of the local transmission dynamics remain poorly quantified, despite the intensive surveillance efforts on the island. We remain uncertain about within-island connectivity due to mosquito mobility. We remain uncertain about the importance of visitors to Bioko Island from mainland Equatorial Guinea in contributing to sustained malaria transmission. MIS data represent a cross-sectional sample of island residents collected during an eight-week period in August and September. In the absence of year-round data, we have ignored seasonal patterns in travel behavior and transmission. The features of the malaria transmission model itself have been simplified: we stick to treating human hosts as either infected or not, and do not model parasitemia, infection history, or changing immune responses of infected hosts. To assess robustness more fully, we would need to vary assumptions about each aspect of the dynamics to know whether it would have a strong effect on the outcome. For this investigation in this particular setting we believe that these simplifying assumptions are unlikely to greatly impact the quantitative results: there is already considerable evidence that many of the cases observed on the island can be attributable to the high endemic level of malaria on the mainland [6, 9, 16, 17]. Ultimately, it may not be possible to eliminate malaria on Bioko Island unless it is part of a regionally coordinated effort.

The style of analysis presented here may be applied to other transmission settings where malaria prevalence is driven in part by non-local exposure and cases imported from other locations. There are a variety of other settings where studies have shown that accounting for non-local exposure and importations is important for understanding the persistence of malaria, including other islands such as Zanzibar [5]; low-transmission regions of sub-Saharan Africa where travel from high-transmission region has been shown to be the primary risk factor for infection [7]; or cases occurring among laborers who become exposed to malaria while working in the forest [8]. The analysis requires spatial data describing malaria prevalence, such as the maps provided by the Malaria Atlas Project, in conjunction with a data set describing travel patterns and behavior. The travel data set may be derived directly from survey data [27], or indirectly inferred from mobile phone data [4], but in any case must make it possible to characterize the duration and intensity of exposure experienced by travelers in the locations they travel to. Together, in conjunction with a mathematical model of malaria transmission, it becomes possible to assess malaria connectivity and quantify how imported cases contribute to local malaria burden.

## Data Availability

All supporting data referred to in the manuscript may be found in a linked Figshare repository

https://doi.org/10.6084/m9.figshare.14380565.v1

## Availability of data and materials

All code required to reproduce the simulations and analysis presented here may be found in the following GitHub repository: https://github.com/dtcitron/bioko_island_travel_materials/releases/tag/v1.0. All data required to parameterize and calibrate the simulations may be found in the following data repository: https://figshare.com/articles/dataset/Data_Supporting_Bioko_Island_Travel_Modeling/14380565

## Author’s contributions

DTC analyzed the data, constructed the simulations, performed the analysis; CAG and GAG provided the survey data from BIMEP; SLW developed the simulation software; SYK, KEB, and HSG provided the geostatistical estimates of malaria prevalence based upon the BIMEP data; DLS developed the modeling methodology.

## Acknowledgements

We acknowledge support from the Bill & Melinda Gates Foundation, Grant OPP1110495. Additionally, we thank the participants of Bioko Island who have taken part in the surveys from which the Bioko data were drawn, the BIMEP field surveyors and supervisors who collected these data, as well as the National Malaria Control Program and the Ministry of Health and Social Welfare of Equatorial Guinea, Marathon Oil, Noble Energy, AMPCO (Atlantic Methanol Production Company), and the Ministry of Mines and Energy of Equatorial Guinea for their continued support of this work. Lastly, we would like to thank the following collaborators for their helpful insights and discussion: Su Yun Kang; Dianna EB Hergott; Olivier Tresor Donfack; Immo Kleinschmidt; Caitlin A Bever; Joshua Suresh; Prashanth Selvaraj; Zhanhao Zhang, John M Henry; Héctor M Sánchez C; Andrew J Dolgert; David S Galick.

## Additional Files

Supplementary Information

