## Supplementary Information for "Quantifying malaria acquired during travel and its role in malaria elimination on Bioko Island"

### 1 Simulation Software Description

To perform the analysis presented in the main text, we developed a stochastic mechanistic simulation of malaria disease dynamics. This simulation software is designed to incorporate data sources relevant to the study of malaria transmission and enable evaluating various anti-malaria intervention policy options.

The simulation model couples a stochastic, agent-based, event-driven model of malaria infection and travel behavior for humans together with a deterministic model of the mosquito vector population dynamics styled after the Ross-Macdonald model [1, 2]. Spatial dynamics are represented through a metapopulation network of patches, where each patch represents an isolated geographical location [3]. Each individual human host is assigned a home patch and may take trips which begin and end at home, visiting another patch for some amount of time. The frequency of trips; the preferred trip destinations; and duration of stay are input as parameters governing the movement behavior of individual human hosts. Infection dynamics in the human population follow Susceptible-Infectious-Susceptible (SIS) dynamics, with elaborations to allow for drug treatment and vaccination. Mosquito populations are modeled much more coarsely as continuous densities, following Susceptible-Exposed-Infectious (SEI) dynamics. While the model allows for mosquitoes to move between patches according to a diffusion matrix, for the present analysis we did not assume mosquitoes diffused between patches.

We have developed a software package called `macro.pfsi` to implement this model. The software package containing the simulation code may be found in the `macro.pfsi` directory at `https://github.com/dtcitron/bioko_island_travel_materials`. The software is an R package which provides a convenient interface to the `C++` simulation algorithm. To be compiled and installed, the package requires a `C++14` compatible compiler. `C++14` features are required to implement event-driven updating of agents using lambda functions as callbacks. Once compiled, `macro.pfsi` can be loaded and called like any other R package. The simulation model writes output to .csv files which can be loaded into R for further analysis and visualization. Our simulation code depends on `Rcpp` to link the compiled code to R, and `RcppArmadillo` for efficient evaluation of the mosquito difference equations and sparse matrix routines to simulate mosquito diffusion. Several vignettes are provided with the package to briefly demonstrate how to parameterize, run, and analyze results of `macro.pfsi`, and all exported functions are documented.

The coupled mosquito-human model dynamics are of SEI-SIS type, common in Ross-Macdonald style models [4]. Because humans are represented as individual agents, rather than an aggregated count of individuals sharing a common state, the software supports individual level heterogeneity in propensity to be bitten by mosquitoes and personal travel habits. Each human agent must be assigned a home patch, which is where they will return to after every trip away from home. Humans also must be assigned a `trip_frequency` parameter ($\pi_{i,j}$ in the mathematical notation presented in Section 4 of this document, where $i$ is the home patch and $j$ is the destination patch), which describes the (Exponentially distributed) frequency at which they choose to take a trip away from home. They also require a vector, `trip_duration` ($\tau_{i,j}$), giving the mean of the exponentially-distributed duration of the trip, which should be equal to the number of patches to allow heterogeneity in duration of stay at different destination patches. Currently, the probability vector for the multinomial distributed choice of trip destination cannot vary by individuals, and is the same for all individuals with the same home patch (it is specified as a probability matrix, $\{\eta_{i,j}\}$).

Each human agent holds an event queue, a list of events sorted by time to fire (occur), such that the soonest event is first. Events may fire in continuous time, and when an event fires a callback function is allowed to change the individual's state, as well as add to queue, alter, or delete future events (each distinct event is denoted by italics). The *infectious bite* event is added to an individual's queue when they receive a bite from an infectious mosquito; upon firing it samples a Bernoulli random variate to determine if that infectious bite will queue an *infection* event after a latent period, which occurs with probability $b$ (transmission efficiency), the individual's probability of infection given an infectious bite. When an infection event fires, a *recovery* event is queued after an exponentially-distributed time to clearance. Additionally, a Bernoulli random variate is sampled to determine if a *fever* event fires after a short delay. Fevers in turn queue *treatment* events, which occur after another short delay. Treatment events cause the individual to clear all parasites and enter a period of prophylaxis where they may not be infected, which decays after a delay (*end prophylaxis* event). Vaccination is simulated by adding a *vaccination* event to the person's queue. Vaccine efficacy is modeled as "all or nothing", being sampled from a Bernoulli random variate. If protective to that individual, the vaccine lowers that individual's $b$ parameter ($b \rightarrow b_{\text{vaxx}} = b/2$), before decaying after a delay (*vaccine wane* event). Treatment may or may not accompany vaccination.

Mosquito dynamics are simulated at a much coarser resolution. The mosquito state for each patch is represented by scalar values $M$ (the total adult female population), $Y$ (the infected adult female population), and $Z$ (the infectious adult female population). The simulation must also keep track of a lagged vector of incubating mosquitoes to account for the extrinsic incubation period (EIP) in the mosquito. The mosquito dynamics are updated according to a discrete daily time step. At the start, a Poisson random variate is sampled for each patch to determine the number of newly emerging adult female mosquitoes (mean emergence rate $\lambda$), which is added to $M$. Then, the force of infection on mosquitoes is calculated as $a\kappa$, where $a$ is the human biting rate, and $\kappa$ is the net infectiousness of humans in each patch (the probability a mosquito becomes infected when blood feeding on *any* human, defined in Eq. 4 below), and $(a\kappa)(M-Y)$ mosquitoes become infected each time step. Those newly infected mosquitoes are added to the vector of incubating mosquitoes. Then, all mosquitoes suffer a constant mortality, diffusion-based movement is simulated (if specified), incubating mosquitoes are shifted up by one day, and those mosquitoes that survive and complete the EIP are added to $Z$. With the exception of stochastic emergence, the mosquito model is equivalent to a discrete time implementation of Ross-Macdonald dynamics with a fixed duration of EIP.

We have described the separate human and mosquito components of the model, but not yet how feedback between them (arising from pathogens being transferred between the two host species) is simulated, which occurs each time step. First, $\kappa$ is computed for each patch. This is done by summing each individual's probability to infect a susceptible mosquito $c$ (transmission efficiency), which changes based on infection status, multiplied by their individual biting heterogeneity weight $w$, and then normalizing by the sum of biting weights. Therefore $\kappa = \frac{\sum_h c_h w_h}{\sum_h w_h}$, where $h \in \mathcal{H}$ are individuals within that patch's population. (In the present analysis, we assume that all biting weights are equal across all human hosts.) Next, each individual computes their individual EIR (entomological inoculation rate). For a patch, the total number of infectious bites arising from the mosquito population is $aZ$, and individual $h$ receives a proportion $\frac{w_h}{\sum_h w_h}$ of those bites, and their product gives the individual level EIR for that person. Because over a small time step, which we choose to be one day, $\kappa$ and EIR do not change significantly, we assume that the mosquito and human populations are conditionally independent given those values calculated at the beginning of the time step. The mosquito state is then updated according to the description given in the previous paragraph, and the value of $\kappa$ just calculated. Then, using their individual value of EIR, each human samples a number of *infectious bite* events that occur over this day and adds them to their event queue. Then, each human fires the next event to happen in their event queue, until the next event would happen on the next time step (day). The algorithm then returns to the top of the loop until the simulation reaches the desired maximum number of time steps (days). A pseudocode description of the algorithm is below.

**macro.pfsi simulation pseudocode algorithm**

```
while(tnow < tmax){

    update_kappa(); // compute net infectiousness of humans
    update_EIR(); // compute EIR for each human

    // update mosquito state
    simulate_mosquito();

    // use EIR to sample infectious bites for each person
    queue_infectious_bites();

    for(h in human_population){
        while(h->next_event_time < tnow){
            fire_next_event(h);
        }
    }

    tnow += 1;
}
```

In the language of stochastic simulation, the model is a hybrid state simulation [5], because mosquito populations are represented as continuous densities and human agents have discrete states they belong to. Mosquito populations update according to a deterministic daily difference equation after stochastic emergence, and humans update according to a continuous-time event driven simulation (due to the presence of some non-Exponential waiting times, it is not a continuous-time Markov chain, but rather a semi-Markov process). Despite elaborations in the human component, when coupled, the dynamics closely fulfill classic Ross-Macdonald assumptions.

### 2 Model Calibration

We calibrate our simulation model by taking inspiration from the source-sink dynamics analysis for the Ross-Macdonald model described in [6, 7]. The simulation software is built to replicate Ross-Macdonald dynamics and we base our calibration off of the standard equilibrium analysis of a set of equations which are constructed to reproduce the Ross-Macdonald dynamics of the simulation software.

We model the disease states of human hosts as follows:

$$
\begin{aligned}
\frac{dX_i}{dt} &= \rho \sum_j \Psi_{i,j} h_j \left(N_i - X_i - P_i\right) - rX_i \\
\frac{dP_i}{dt} &= (1-\rho) \sum_j \Psi_{i,j} h_j \left(N_i - X_i - P_i\right) - \eta P_i
\end{aligned}
\tag{1}
$$

The matrix element $\Psi_{i,j}$ represents the fraction of time that a resident of patch $i$ spends in patch $j$. We are able to derive an the matrix $\Psi$ using the movement parameters discussed in the Section 3 below. Using the parameters representing the rates of traveling $\phi_{i,j}$ and returning $\tau_{i,j}$, we can write out a set of equations which describe how many people from location $i$ are found either at home (location $i$) or away (location $j$).

$$\frac{dN_{i,i}}{dt} = -\sum_{j=1}^{K} \phi_{i,j} N_{i,i} + \sum_{j=1}^{K} \tau_{i,j} N_{i,j}$$
$$\frac{dN_{i,j}}{dt} = -\tau_{i,j} N_{i,j} + \phi_{i,j} N_{i,i}$$

Solving for the equilibrium states, we can obtain expressions $\Psi_{i,j} = N_{i,j}/N_i$:

$$\Psi^*_{i,i} = \frac{1}{1 + \sum_{k=1}^{K} \frac{\phi_{i,k}}{\tau_{i,k}}}$$
$$\Psi^*_{i,j} = \frac{\phi_{i,j}}{\tau_{i,j}} \frac{1}{1 + \sum_{k=1}^{K} \frac{\phi_{i,k}}{\tau_{i,k}}} \qquad i \neq j$$

The constant $h_j$ in Eq. 1 represents the force of infection (FOI) experienced by a user in location $j$. We use a standard equilibrium analysis approach to Eq. 1 and set the left-hand sides to zero, yielding an expression for the FOI in terms of prevalence $X_i/N_i$:

$$\sum_j \Psi_{i,j} h_j = \frac{r X_i/N_i}{(1-\rho)\left(1 - \left(1 + \frac{r\rho}{\eta(1-\rho)} \frac{X_i}{N_i}\right)\right)} \tag{2}$$

To calibrate the simulation model, we use the mapped estimates of prevalence from [8] along with equation 2 to obtain the FOI in each patch. In practice, we obtain the vector of values for $h_j$ by inverting the $\Psi$ matrix and multiplying it by the right side of Eq. 2. We note that over the course of calibrating thousands of simulations based on different $Pf$PR surfaces that there are a handful of instances where the resulting FOI in a small number of patches runs negative. These patches turn out to be areas with very few residents and very low $Pf$PR, where there is no way to numerically reconcile the low $Pf$PR with the high rate of importation from the travel model. In these rare cases, we set the local FOI to zero. Because these patches tend to have fewer than 10 residents, this correction does not affect the large majority of the simulation and does not impact the analysis.

The next step in calibration is relating the FOI ($h$) of each patch back to the mosquito activity in the simulation model. The Ross-Macdonald dynamics define the FOI as arising from the interactions between infectious mosquitoes and available human hosts:

$$h_i = abZ_i / \sum_j \Psi_{j,i} N_j = \frac{M_i}{\sum_j \Psi_{j,i} N_j} ab \frac{Z_i}{M_i} \tag{3}$$

We model mosquito population dynamics and disease states as follows:

$$\begin{aligned} \frac{dZ_i}{dt} &= ac\kappa_i \left( M_i e^{-gn} - Z_i \right) - gZ_i \\ \kappa_i &= \frac{\sum_j \Psi_{j,i} X_j}{\sum_j \Psi_{j,i} N_j} \end{aligned} \tag{4}$$

The expression $\kappa_i$ represents the fraction of infected human hosts located at location $i$, using $\Psi$ to account for the fact that human hosts are allowed to travel between patches. We again use standard equilibrium analysis and solve for the fraction of infectious mosquitoes:

$$\frac{Z_i}{M_i} = \frac{ac\kappa_i e^{-gn}}{ac\kappa_i + g}$$

The above expression, combined with the expression for FOI in Eq. 3, makes it possible to solve for the total adult mosquito population $M_i$ in each patch at equilibrium. The last step is to calibrate the emergence rate $\lambda_i$ in each patch such that the equilibrium adult mosquito population matches $M_i$.

These calibration steps may be found written out as code in lines 81-128 in the simulation script found in `simulation_configurations/baseline_simulation_configuration.R` at `https://github.com/dtcitron/bioko_island_travel_materials/releases/tag/v1.0`.

### 3 Movement Model Parameterization from Data

The high quality travel survey data made available through the Bioko Island Malaria Elimination Program (BIMEP) Malaria Indicator Surveys (MIS) provides an unprecedented opportunity to construct and parameterize a detailed model of human host travel patterns. As discussed in the main text, we model each human host's travel behavior in three steps: the human host chooses when to leave their home patch, chooses the destination, and chooses how many days they spend away before returning to their home patch. Within the simulation, each individual requires a set of parameters that includes the frequency of travel; the multinomial probability distribution for determining travel destination; and the mean duration of their trip [9]. We use the MIS data from 2015 through 2018 to parameterize each of these sets of parameters [10, 11, 12, 13].

Using mathematical notation, we represent the overall rate a which a user travels $i \to j$ as $\phi_{i,j}$, and break up that rate into a product of the rate at which a user in location $i$ leaves home ($\pi_i$) times the probability of choosing to travel to $j$ given one's home residence location at $i$ ($\eta_{i,j} \equiv \mathbf{P}(i \to j \mid i)$):

$$\phi_{i,j} = \pi_i \times \eta_{i,j}$$

#### Frequency of Leaving Home

Code supporting this section is available online in `Trip_frequency_model.R` in the accompanying GitHub repo `https://github.com/dtcitron/bioko_island_travel_materials/releases/tag/v1.0`.

Respondents in the MIS reported whether they had traveled within the last 60 days. From this, we can model the probability of whether an average survey respondent left the island using a binomial probability model. As covariates, we account for the local population of each respondent's residence patch; the Administrative subdivision containing their residence patch; and the distance of the home residence patch from the centroid of Malabo district (that is to say, the distance from the city). For Administrative subdivisions of the home residences, we used the six subdivisions from the survey (Malabo, Baney, Luba, Riaba, Moka, Ureka) as well as one extra category used for patches located in Malabo district but outside of the urban center (peri-urban Malabo) — this latter distinction was important for improving the model fit as it differentiates the movement patterns of those who live within and without the city.

Fitting the statistical model to data, we can make estimates of the probability that a respondent leaves home in the 60 day period. These estimates are available everywhere, even in locations without survey responses. We show a map of the probability of leaving home in Figure 1, noting how the frequency of leaving home increases as one gets further south on the island. Within the simulation, we use these probabilities to parameterize the average rate of leaving home ($\pi_i$) by dividing the probability of leaving home by the duration of the study period (60 days).

#### Destination Choice Probability Model

Code supporting this section is available online in `Trip_destination_choice_model.R` and `region_to_areaId_mapping.R` in the accompanying GitHub repo `https://github.com/dtcitron/bioko_island_travel_materials/releases/tag/v1.0`.

Respondents in the MIS who reported travel within the last 60 days also reported which Administrative subdivision they had traveled to. They would indicate the destination as one of Malabo, Baney, Luba, Riaba, Moka, Ureka, or Off-Island. For on-island travel, this is a source of additional ambiguity which we must address: if someone reported that they traveled to one of these destination regions, which specific patch did they travel to? We adjust our destination choice model to represent travelers first choosing a destination region $d$ and then choosing a specific patch $j$ within that destination region:

$$\eta_{i,j} \equiv \mathbf{P}(i \to j \mid i) = \mathbf{P}(i \to d \mid i) \times \mathbf{P}(d \to j \mid i, d)$$

We model the first term $\mathbf{P}(i \to d \mid i)$ by first trying to estimate the mean number of trips taken by all patch residents from each patch to each of the seven destination regions. We model the on-island trips and off-island trips separately, seeing as traveling off-island requires extra effort and cost on the part of the traveler (e.g. a boat or a plane). We use a negative binomial probability model to predict the trip counts. For patch covariates, we take inspiration from gravity models of movement [14, 15] and allow our model to depend on the population of the origin; the population of the destination; the distance between the origin and the destination; the region where the origin patch is located (again differentiating between urban and peri-urban Malabo); and the distance from the origin to the centroid of Malabo district. Taking inspiration from [16], we use a log-transformed origin-destination distance

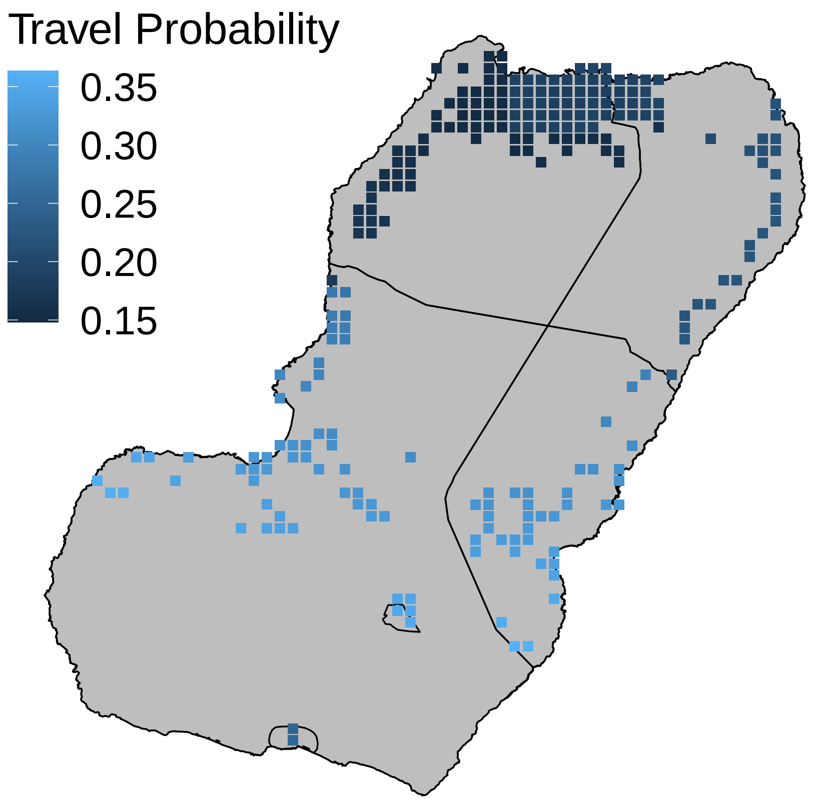


**Figure 1** Modeled estimates of probability that an average resident leaves home within the 60-day period.

to inform the off-island travel model but not for the on-island travel model. Using this model to predict the mean number of trips $i \to d$, we can divide out by the total number of trips to obtain the normalized multinomial probability distribution for each patch ($\mathbf{P}(i \to d \mid i)$).

A few examples are shown in Figure 2 below. From the destination choice model, we find that residents of Malabo are most likely to travel off-island compared with the residents of other regions. Residents in the southern parts of the island prefer to travel to Malabo in the north.

We have no data to directly inform which exact patch a user may have traveled to within any given region. To model the second term $\mathbf{P}(d \to j \mid i, d)$, we remain agnostic and assume that the average traveler has an equal probability of traveling to visit any resident of the destination region they go to. That is to say, within each destination region, we weight the probability of traveling to patch $j$ according to the population in patch $j$. Changing this assumption to instead weight all patches equally (equal probability of visiting any patch within the region) does not impact the results discussed in the main text.

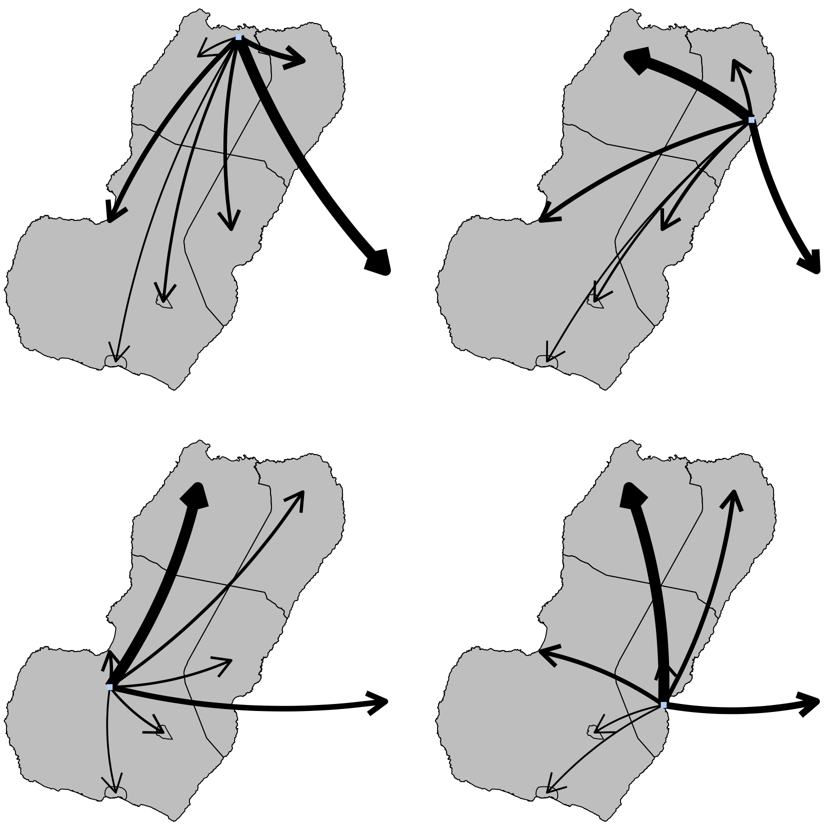

**Figure 2** Diagram showing relative probability weights of traveling to each destination from four example locations on Bioko Island.

#### Trip Duration

Code supporting this section is available online in `Trip_duration_model.R` in the accompanying GitHub repo `https://github.com/dtcitron/bioko_island_travel_materials/releases/tag/v1.0`.

The 2018 MIS included a question on the duration of travel [13], which proved to be crucial for understanding of how long travelers were exposed while away from their homes. Figure 3 shows histograms of the reported number of days away from home, for both off-island (left) and on-island (right) trips.

We use an exponential waiting time model to estimate the mean duration of travel for each category of trip. For off-island travel, we estimate a mean travel duration of 21.2 days. For on-island travel, we estimate a mean travel duration of 10.3 days. Inverting this, we obtain constant rates for returning home from travel which depend on the trip destination location.

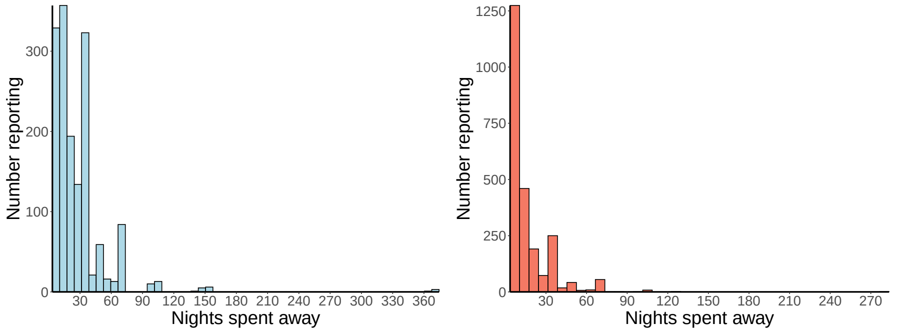


**Figure 3** **Left**: Histogram of trip durations from Bioko Island to mainland Equatorial Guinea. **Right**: Histogram of durations of trips within Bioko Island.

### 4 Mathematical Notation and Definitions

**Table 1** Movement Model Parameter Definitions

| Parameter | Interpretation |
|---|---|
| $\phi_{i,j}$ | Rate at which a resident of $i$ travels $i \to j$ |
| $\pi_i$ | Rate at which a resident of $i$ leaves home |
| $\eta_{i,j}$ | Destination choice probability distribution choosing destination location $j$ given home at $i$ |
| $\tau_{i,j}$ | Rate at which a visitor to $j$ returns home to $i$ |
| $\Psi_{i,j}$ | Average fraction of time spent in location $j$ for residents of location $i$ |

**Table 2** State Variables and Parameters

| Variable or Parameter | Interpretation | Value (if constant) |
|---|---|---|
| $N$ | Total number of humans | Calibrated from census |
| $X$ | Number of infected and infectious human hosts | |
| $P$ | Number of treated/protected human hosts | |
| $\lambda$ | Adult mosquito emergence rate | Indirectly calibrated from MAP data [17] |
| $M$ | Total number of mosquitoes | |
| $Y$ | Number of infected but not yet infectious mosquitoes | |
| $Z$ | Number of infectious mosquitoes | Surviving EIP |
| $r$ | Rate of human host recovery | $r = 1/200$ day$^{-1}$ |
| $\rho$ | Probability of receiving treatment after infection | $\rho = 0.067$ |
| $\eta$ | Rate at which treatment effects wear off | $\eta = 1/32$ day$^{-1}$ |
| $a$ | Mosquito bites per human per day | $a = 0.27$ |
| $b$ | Mosquito-to-human transmission efficiency | $b = 0.55$ |
| $c$ | Human-to-mosquito transmission efficiency | $c = 0.15$ |
| $g$ | Rate of mosquito death | $g = 0.1$ |
| $e^{-gn}$ | Fraction of mosquitoes who survive the extrinsic incubation period (EIP) | |
| $b_{\text{vaxx}}$ | Reduced $b$ for a vaccinated individual | $b_{\text{vaxx}} = b/2$ |
| $\mu_{\text{vaxx}}$ | Mean duration of vaccination efficacy | $\mu_{\text{vaxx}} = 300$ days |
| $\sigma_{\text{vaxx}}$ | Mean duration of vaccination efficacy | $\sigma_{\text{vaxx}} = 60$ days |

Each of the state variables and parameters listed above in Table 4 are permitted to vary between different patches in the most general multipatch model. In the present study, we allow the state variables human populations and adult mosquito emergence rates to vary across different patches (locations) in the model, but we fix the bionomic parameters ($r, a, b$, etc.) and assume they are constant at all locations.

**Author details**
[1]University of Washington, Institute for Health Metrics and Evaluation, 2301 Fifth Ave., Suite 600, 98121 Seattle, WA, USA. [2]Medical Care Development International, 8401 Colesville Road, Suite 425, 20910 Silver Spring, MD, USA. [3]Medical Care Development International, 8401 Colesville Road, Suite 425, 20910 Silver Spring, MD, USA. [4]University of Oxford, Malaria Atlas Project, Big Data Institute, Li Ka Shing Centre for Health Information and Discovery, OX3 7LF Oxford, UK.
